# AI-Driven Feature Selection Using Only Survey Variable Descriptions: Large Language Models Identify Adolescent Vaping Predictors

**DOI:** 10.64898/2026.03.06.26347816

**Authors:** Kaidi Zhang, Zimo Zhao, Yan Hu, Thuy T. T. Le

**Author notes:** Corresponding author: Thuy T. T. Le, PhD, Department of Health Management and Policy, School of Public Health, University of Michigan, 1415 Washington Heights, Ann Arbor, MI, 48109, United States. These authors contributed equally to this work (Kaidi Zhang, Zimo Zhao).

## Abstract

**Objective:** To evaluate the effectiveness of various Large Language Models (LLMs) in identifying reliable predictors of Electronic Nicotine Delivery Systems (ENDS) initiation among adolescents, using solely large-scale survey variable descriptions.

**Methods:** A cohort of 7,943 tobacco-naïve adolescents aged 12-16 years from the Population Assessment of Tobacco and Health (PATH) Study was analyzed to predict ENDS use at wave 5. Four instruction-tuned LLMs – GPT-4o, LLaMA 3.1-70B, Qwen 2.5-72B-Instruct, and DeepSeek-V3 – were systematically evaluated for text-based feature selection using only variable descriptions from wave 4.5. Selected features were used to train LightGBM classifiers, with model performance compared to a baseline.

**Results:** Our findings reveal notable consistency among the four instruction-tuned LLMs, with substantial overlap in the top predictors each model identified. These selected variables spanned critical domains such as peer and household influence, risk perception, and exposure to tobacco-related cues. LightGBM classifiers trained on PATH wave 4.5-5 data using features selected by the LLMs demonstrated strong predictive performance. Notably, Qwen 2.5-72B-Instruct achieved an AUC of 0.791 with 30 predictors, surpassing the baseline AUC of 0.768.

**Discussion:** The substantial overlap among the top predictors identified by different LLMs suggests a shared reasoning process, despite variations in model architecture and training. LightGBM classifiers trained on these LLM-selected features achieved performance comparable to, or exceeding, models trained on the full set of survey variables, underscoring the high quality of features selected solely from textual descriptions. Moreover, these findings are consistent with previous tobacco regulatory research, further validating the effectiveness of LLM-driven feature selection.

**Conclusion:** Instruction-tuned large language models can effectively perform text-based feature selection using survey variable descriptions alone, without accessing raw survey data. This scalable, interpretable, and privacy-preserving framework holds promise for behavioral health research and tobacco use surveillance.

## Introduction

The tobacco landscape has changed significantly over the last decade with the emergence and growing popularity of novel products such as electronic nicotine delivery systems (ENDS), oral nicotine pouches, and heated tobacco products [1]. The diversity of the tobacco market makes it challenging to understand tobacco use behaviors.

In tobacco research, survey data are the primary source of information for studying tobacco use patterns and tobacco-related health effects. Given these massive, high-dimensional datasets, traditional statistical approaches such as regression fall short in leveraging all the available information. This is due to their reliance on standard assumptions and expert knowledge to select adjusted covariates [2]. A growing body of research is focusing on applying advanced machine learning (ML) techniques to better understand tobacco use behaviors [3]. In these studies, researchers have adopted ML methods to extract the most informative predictive features from high-dimensional datasets. Traditional feature selection techniques, such as recursive feature elimination (RFE) [4], mutual information [5], and Lasso regression [6], have been widely used to reduce dimensionality and enhance model performance [7, 8]. Furthermore, SHAP (SHapley Additive exPlanations) is utilized to improve model interpretability by quantifying each feature’s contribution to the model output based on game theory [9]. These methods, often combined with ML models like XGBoost and random forests, have shown promise in public health applications [2, 10]. However, these approaches can be unstable across datasets, require rigorous training on empirical data, and remain sensitive to sample-specific biases.

Several recent studies have explored the use of LLMs to address tobacco-related issues, such as the identification of tobacco products in social media videos, tobacco-promoting social media content, and promoting vaping cessation [11-13]. Latest advances in LLMs have opened new avenues for LLM-based feature selection. Studies have shown that instruction-tuned LLMs can perform zero-shot variable evaluation by leveraging semantic context encoded in textual variable descriptions – without accessing individual-level data [14, 15]. To our knowledge, Le’s work [16] is among the first in tobacco regulatory science to explore the use of an LLM, OpenAI’s GPT-4.1, to perform text-based feature selection for predicting smoking cessation. However, experimenting and comparing the efficacy of multiple LLMs for text-based feature selection remains untouched.

Unlike previous approaches that require iterative retraining and rely on data-driven heuristics, an LLM-guided method provides a lightweight, transparent, and scalable solution to variable screening in complex health surveys. In addition, by averaging importance scores across multiple LLMs and evaluating stability via statistical dispersion metrics (e.g., variance, coefficient of variation), we ensure the consistency of selected features. Empirical results demonstrate that our approach not only improves classification performance over conventional baselines but also highlights non-trivial predictors that may have been overlooked by standard statistical models.

Our findings contribute to the emerging literature on LLM-driven feature engineering [14-16] and offer practical tools for public health researchers seeking to leverage foundation models in real-world epidemiological settings.

## Materials and Methods

### Data

Data were obtained from the PATH Study – a nationally representative, longitudinal cohort study assessing tobacco use and associated health outcomes in the United States [17]. This analysis included merged data from youth waves 4.5 (December 2017 to December 2018) and 5 (December 2018 to November 2019), as the use of ENDS increased most significantly in these groups during this period [18]. Our baseline population included tobacco-naive participants, who aged from 12-16 years and had never tried any tobacco products, including smoked cigarettes, traditional cigars, cigarillos, filtered cigars, pipe, hookah, bidis, or kretek, and had never used e-products, smokeless tobacco, snus pouches, or dissolvable tobacco in wave 4.5. The outcome of interest is their ENDS use status at wave 5, which was assessed with a binary response format (1 = yes, 2 = no), indicating whether the participant had used ENDS in the past 30 days. Informed consent was waived because this work analyzed a de-identified publicly available dataset.

After merging the data from wave 4.5 with the outcome variable, ENDS use status, from wave 5, and extracting a subpopulation of tobacco-naïve 12-16-year-olds, we obtained a dataset with 7966 participants and 1396 survey variables spanning multiple domains, including personal health status, family relationships, parental characteristics, and social network influences. We then removed irrelevant variables, variables with more than 2.5% missing values or low variation, and individuals with missing wave 5 ENDS use status. As a result, we obtained a dataset with 7943 adolescents and 215 wave 4.5 variables. Additionally, we excluded participants with any missing values in key demographic or behavioral baseline variables.

After data preprocessing, we applied four commercially available LLMs – GPT-4o, LLaMA 3.1-70B, Qwen 2.5-72B-Instruct, and DeepSeek-V3 – to identify the top variables most predictive of adolescent ENDS use, selecting sets of 50 down to 10 variables in decrements of five. These models performed feature selection based solely on the textual descriptions of survey variables, without direct access to the actual data. To assess the potential bias introduced by model-specific reasoning, we conducted statistical comparisons across the variable importance scores generated by each LLM. After feature selection, we removed individuals with any missing data in the selected top variables, thereby retaining as many valuable samples as possible. The specific sample size for each set of selected top variables is shown in Table 1. For comparison, removing individuals with any missing data across all 214 wave 4.5 variables would result in a dataset of 5,996 participants. For each set of selected top variables, the final clean dataset was split into a training set (80%), which was used to fine-tune model hyperparameters and train LightGBM for predicting ENDS use status, and a hold-out test set (20%) used solely for evaluating model performance on unseen data.

**Table 1.** Details of the filtered data. Values in parentheses represent the number of individuals who reported current ENDS use in wave 5.

| Model | 50F | 45F | 40F | 35F | 30F | 25F | 20F | 15F | 10F |
| --- | --- | --- | --- | --- | --- | --- | --- | --- | --- |
| **LLaMA 3.1-70B** | 6815 | 6835 | 6848 | 6876 | 6980 | 7041 | 7154 | 7275 | 7383 |
|  | (295) | (296) | (298) | (298) | (302) | (305) | (309) | (313) | (318) |
| **Qwen 2.5-72B-Instruct** | 6799 | 6820 | 6974 | 7056 | 7081 | 7174 | 7248 | 7367 | 7487 |
|  | (295) | (295) | (302) | (303) | (305) | (307) | (312) | (317) | (319) |
| **GPT-4o** | 6862 | 6889 | 6937 | 6961 | 6983 | 7099 | 7157 | 7297 | 7433 |
|  | (297) | (298) | (298) | (299) | (300) | (305) | (310) | (312) | (320) |
| **DeepSeek-V3** | 6789 | 6829 | 6912 | 6975 | 7071 | 7157 | 7208 | 7389 | 7538 |
|  | (293) | (296) | (300) | (304) | (304) | (310) | (311) | (316) | (320) |

### Text Driven Feature Selection

To improve the accuracy of ML models and conserve computational resources, while investigating the feasibility of employing LLMs for feature selection, we conducted experiments to select the most relevant variables from wave 4.5 for predicting adolescent ENDS use. In this experiment, we utilized four state-of-the-art instruction-tuned LLMs – GPT-4o [19], LLaMA 3.1-70B [20], Qwen 2.5-72B-Instruct [21], and DeepSeek-V3 [22], which were prompted to evaluate the relevance of each baseline variable (a total of 214 wave 4.5 variables) for predicting future ENDS use. Specifically, we presented each variable name and its description to the LLMs and asked the models to assign an importance score between 0 and 1 to each variable, with 1 indicating the strongest influence on predicting ENDS use status. We repeated this procedure for each of the four LLMs across 15 independent runs. For each model, we then computed the mean importance score for each variable by averaging the scores obtained from the 15 runs. Further details on our prompt design are described in the Appendix. We set the parameters temperature, top p, and top k to 0.7, 0.7, and 50, respectively, to achieve finer control over the randomness and diversity of the generated text, balancing creativity and stability through their combined effect. This configuration ensures that the LLMs can robustly and comprehensively analyze the importance of various factors in predicting ENDS use status.

### Utilizing LightGBM and LLM-Selected Variables to Predict ENDS Use Status

LightGBM is an efficient gradient boosting decision tree framework that achieves fast training and high-precision predictions on datasets through histogram acceleration techniques, Gradient-based One-Side Sampling, and Exclusive Feature Bundling [23]. Known for its low computational cost and strong handling of sparse data, LightGBM has demonstrated excellent performance on tasks such as classification and regression, and is popular in the ML community [24-26]. For each LLM, we ranked the variables based on the mean scores to select the top 50 variables with the highest scores (referred to as the top 50 variables). Then, we started with training LightGBM on the training dataset with the top 50 variables, and then gradually reduced the number of variables to test the effectiveness of our variable selection. Specifically, the number of variables used in the experiments was set to 50, 45, 40, 35, 30, 25, 20, 15, and 10. We used parallel computing, aiming to improve the computing speed and the generalization ability of the model. To be specific, the model training within each Optuna hyperparameter trial uses a fixed number of threads THREADS PER TRIAL = 8 and is enforced through the LightGBM parameter num threads = THREADS PER TRIAL. Meanwhile, we set the environment variables OMP NUM THREADS and MKL NUM THREADS to the same value to ensure that the underlying OpenMP/MKL library only occupies this part of the threads during data preprocessing and matrix operations. Optuna maximized the mean cross-validated area under the ROC curve (CV-AUC) [27]. The specific hyperparameters are shown in Table 2. Optuna is an open-source automated hyperparameter optimization framework that adopts an efficient Bayesian optimization algorithm and supports functions such as distributed optimization and dynamic search space [28]. It is widely used in the automatic parameter tuning tasks of ML and deep learning models.

**Table 2.**
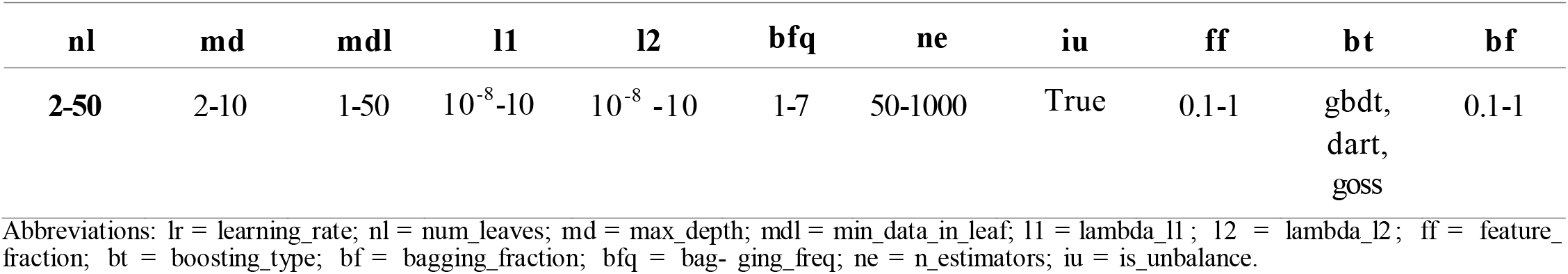
Hyperparameter search space explored by Optuna.

We trained the model on the training data with 5-fold cross-validation and then evaluated its performance on the test data. Each trial trained LightGBM [23] enhanced with Gradient-based One-Side Sampling and Dropouts meets Multiple Additive Regression Trees for up to 1000 boosting rounds, with early stopping after 50 non-improving iterations. Class imbalance was addressed by setting is unbalance to True, which internally up-weights minority-class observations. For a specific number of predictors generated by one of the four LLMs, we trained LightGBM 100 times with random seeds ranging from 1 to 100 (all other settings remain the same) and computed the mean AUC.

## Results

Figure 1 shows the stability metrics, including relative mean deviation (RMD), coefficient of variation (CV), and variance, for the scores generated by DeepSeek-V3, LLaMA 3.1-70B, GPT-4o, and Qwen-72B-Instruct. For each LLM, the importance scores assigned to the same variables remained stable across 15 independent runs, indicating strong internal consistency. Specifically, across all models, the RMD ranges from 0 to 0.15, the variance ranges from 0 to 0.018, and the CV ranges from 0 to 0.12. The four lists of the top 50 variables generated by the LLMs shared 31 variables in common; see Table S5 in the Appendix for details.

**Figure 1.**
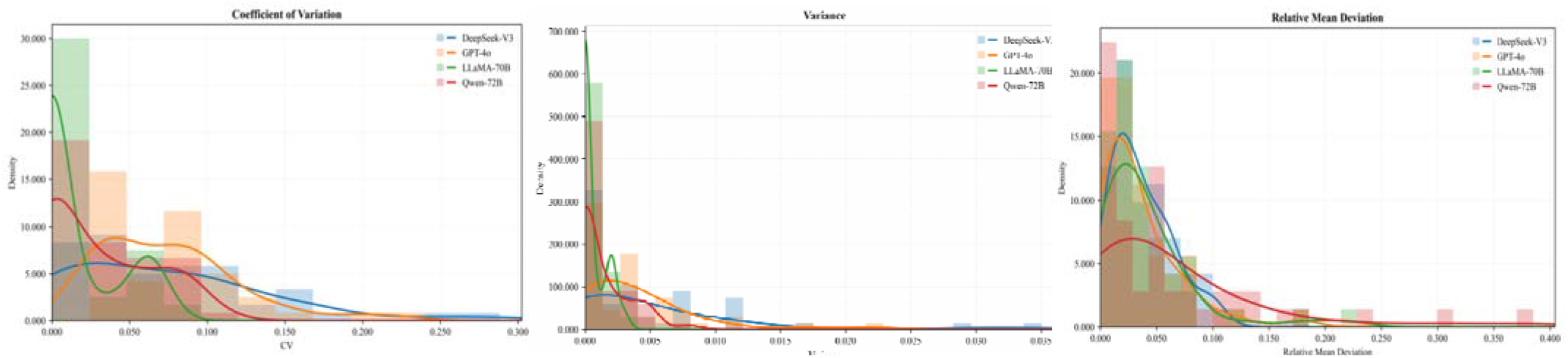
Stability metrics for feature importance scores generated by four large language models (LLMs).

LightGBM trained with all 214 variables yielded an average AUC of 0.768 (standard deviation (SD): 0.027). Across all four LLMs, LightGBM consistently achieved high predictive performance with reduced feature sets, comparable to its performance using all available variables. When selecting 30 features, the Qwen-72B-Instruct model achieved the highest average AUC (mean: 0.791, SD: 0.024). For DeepSeek-V3, the highest average AUC (mean: 0.772, SD: 0.032) was obtained when selecting 35 features. Regarding GPT-4o, the model reached its highest average AUC (mean: 0.784, SD: 0.027) when using 35 features. LLaMA 3.1-70B performed notably well with 40 features, achieving an average AUC of 0.789 (SD: 0.029). As shown in Table 3, the performance of LightGBM using the selected variable subsets is comparable to the model’s performance when utilizing all 214 variables.

**Table 3.** Mean (standard deviation) of AUC achieved by LightGBM for various feature set sizes. Standard deviation values are shown in parentheses.

| Features | All variables | Qwen-72B-Instruct | DeepSeek-V3 | GPT-4o | LLaMA 3.1-70B |
| --- | --- | --- | --- | --- | --- |
| **214** | 0.768 (0.027) |  |  |  |  |
| **50** |  | 0.777 (0.033) | 0.763 (0.024) | 0.781 (0.026) | 0.777 (0.029) |
| **45** |  | 0.781 (0.026) | 0.752 (0.033) | 0.771 (0.033) | 0.787 (0.030) |
| **40** |  | 0.776 (0.036) | 0.763 (0.037) | 0.778 (0.033) | **0.789 (0.029)** |
| **35** |  | 0.784 (0.034) | **0.772 (0.032)** | **0.784 (0.027)** | 0.784 (0.035) |
| **30** |  | **0.791 (0.024)** | 0.755 (0.035) | 0.775 (0.034) | 0.779 (0.022) |
| **25** |  | 0.789 (0.029) | 0.765 (0.028) | 0.764 (0.033) | 0.787 (0.023) |
| **20** |  | 0.767 (0.041) | 0.752 (0.016) | 0.766 (0.025) | 0.761 (0.026) |
| **15** |  | 0.759 (0.038) | 0.741 (0.007) | 0.733 (0.036) | 0.741 (0.031) |
| **10** |  | 0.729 (0.025) | 0.748 (0.009) | 0.698 (0.021) | 0.715 (0.031) |

Additionally, the top 10 most meaningful variables selected by various LLMs in predicting Wave 5 ENDS use status are presented in Tables S1-S4 in the Appendix.

## Discussion

This work aimed to evaluate and compare the performance of four different LLMs (DeepSeek-V3, LLaMA 3.1-70B, GPT-4o, and Qwen-72B-Instruct) in selecting the most informative wave 4.5 predictors of wave 5 ENDS use status among tobacco-naive youth aged 12-16 at baseline, using only the descriptions of PATH survey variables. To our knowledge, our study is among the first to systematically explore the efficacy of multiple LLMs for text-based feature selection in tobacco regulatory research.

Using the same prompt design, the lists of the top 50 variables generated by these models show a high degree of overlap, with 31 variables in common. This indicates remarkable consistency in the models’ reasoning processes despite differences in their underlying model architectures and training data. These common variables selected by all four LLMs encompass a range of factors related to family and peer influence, tobacco risk perceptions, tobacco access and exposure, personal attitudes and intentions, as well as exposure to tobacco advertisements and promotions (see Table S5 in the Appendix). In a previous study [10], the author identified the top 44 predictors of future ENDS use status using the same data source but a different methodology; these predictors also reflected similar domains. The consistency between these findings further validates the capability of LLMs to perform effective text-based feature selection. Furthermore, the predictive performance of LightGBM trained on the set of 31 common variables – as well as on the reduced feature sets generated by each LLM – is comparable to, or even surpasses, its performance when trained on all 214 variables (see Table 3). Additionally, it matches the performance of XGBoost trained on the top 44 variables selected based on the highest mean SHAP values using similar data, as reported in Le’s study [10]. The consistently high predictive performance of LightGBM trained on different feature set sizes selected by LLMs further underscores the high quality of these selected variables. Together, the significant overlap among the top 50 variables and the high predictive performance of LightGBM with these selected variables suggest the reliability of instruction-tuned LLMs in performing feature selection based solely on the textual descriptions of variables.

Since the data sample size associated with each set of predictors selected by each LLM varies, as shown in Table 1, the quality and distribution of the corresponding training and test data also differ. Consequently, the high performance of LightGBM with a particular LLM-selected feature set (see Table 3) does not necessarily indicate that the set of predictors chosen by that LLM is superior to those identified by the others. Rather, these performance differences may be influenced by variations in sample size and data quality across the predictor sets.

Embedding LLM-generated semantic priors within ML pipelines offers a potential approach to balance predictive power and interpretability. The approach is model-agnostic, privacy-preserving (no raw data are exposed during scoring), and readily extensible to other high-dimensional public-health datasets.

## Limitations

First, the LLMs evaluated here were not fine-tuned on PATH-specific language; domain adaptation might yield further gains. Second, reliance on a top-k threshold may omit weak but complementary predictors. Future work should explore soft-thresholding or regularization-based integration of LLM scores. Furthermore, since LLM-based feature selection relies heavily on the quality of the survey questionnaire, careful survey design is essential to ensure the high performance of this approach. Like in studies that use LLM-based approaches, our work have other limitations such as biases in pre-trained data, occasional generation of inaccurate information (“hallucinations”), and variability in reproducibility related to prompt sensitivity, and others as described in [16] and references therein. Continued efforts toward model transparency, external validation, and standardized evaluation will help mitigate these challenges and enhance reliability in applied research settings.

## Conclusion

LLM-assisted feature engineering improves both the predictive accuracy and interpretability of e-cigarette use among adolescents. Integrating LLM with other ML methods provides a promising approach for addressing public health research questions using large empirical datasets.

## Data Availability

The PATH Study data used for analysis are accessible to the public.

https://www.icpsr.umich.edu/web/NAHDAP/studies/36498

## Declaration of Competing Interests

The authors report there are no competing interests to declare.

## Funding

No funding was received for this research.

## Notes

### Competing Interest Statement

The authors have declared no competing interest.

### Funding Statement

None

