## Supplemental Material for "AI-Driven Feature Selection Using Only Survey Variable Descriptions: Large Language Models Identify Adolescent Vaping Predictors"

Appendix to the manuscript entitled **“AI-Driven Feature Selection Using Only Survey Variable Descriptions: Large Language Models Identify Adolescent Vaping Predictors”**

Kaidi Zhang^1+^, Zimo Zhao^1+^, Yan Hu^1,2^, Thuy T. T. Le^3*^

^1^School of Data Science, The Chinese University of Hong Kong, Shenzhen, Guangdong 518172, China

^2^National Health Data Institute, Shenzhen, Guangdong 518172, China

^3^Department of Health Management and Policy, School of Public Health, University of Michigan, Ann Arbor,

MI 48109, USA

+These authors contributed equally to this work.

* Corresponding author: Thuy T. T. Le, PhD, Department of Health Management and Policy, School of Public Health, University of Michigan, 1415 Washington Heights, Ann Arbor, MI, 48109, United States. Telephone: +734-764-6036;.

**LLM-based feature-scoring instruction**

This is to present the system- and user-level instructions provided to a large-language model (LLM). This two-tiered prompting system clearly outlines: (i) the study objectives, (ii) the variable scoring tasks, (iii) the epidemiological context, and (iv) the required JSON-formatted outputs. The specific user instruction targeting variable evaluation was systematically generated 206 times, once for each baseline variable assessed at Wave 4.5, resulting in 206 distinct JSON objects. Each variable was then ranked according to the “score” field returned by the LLM. Subsequently, the top-scoring predictors (default k = 50) were selected for further multivariable analysis.

**1. System instruction**

**Study definition**

Identify key variables measured in Wave 4.5 (Dec 2017 - Dec 2018) of the Population Assessment of Tobacco and Health (PATH) Study that are associated with electronic nicotine delivery systems (ENDS) use in Wave 5 (Dec 2018 - Nov 2019) among adolescents who were tobacco-naïve at baseline.

**Scoring task**

For each candidate baseline variable, produce a numerical relevance score between 0 and 1. Higher values indicate a stronger relationship with the target concept. These scores will later be used to select the top k most important predictors (default k = 50).

**Dataset context**

Responses originate from a nationally representative questionnaire survey of U.S. teenagers, including more than 200 variables (e.g., age, gender, parental smoking, risk perception). The primary goal is to identify the subset of baseline factors that best predict future ENDS initiation.

**Required output**

Return valid JSON with the following two keys:

{"reasoning": "Brief justification", "score": <numeric value 0{1>}

**2. User instruction (generated once per variable)**

The target concept is whether a teenager will initiate e-cigarette (ENDS) use in the future. Please provide both a numerical relevance score and a concise justification for the feature “{variable name} ", described as: {variable description}. The output must be valid JSON with two fields:

• reasoning – one to two sentences briefly explaining the epidemiological or behavioral rationale.

• score – a numeric value between 0 and 1 reflecting the estimated relevance. Example JSON

{"reasoning": "Peer ENDS exposure increases initiation risk via social influence.", "score": 0.82}

| **Order** | **Variable** | **Variable description** | **Importance Score** |
| --- | --- | --- | --- |
| 1 | X04 YX0716 | In next 30 days, likely to purchase e-cigarettes or electronic nicotine products | 0.900 |
| 2 | X04 YX0681 | How many of your best friends use e-cigarettes | 0.897 |
| 3 | X04 YX0678 | In past 30 days, received a free sample of an e-cigarette or other electronic nicotine product | 0.893 |
| 4 | X04 YX0715 | Level of ease for youth to buy e-cigarettes or other electronic nicotine products | 0.873 |
| 5 | X04 YX0741 02 | People who are important to you use: E-cigarettes or other electronic nicotine products | 0.873 |
| 6 | X04 YX0709 | Has a favorite e-cigarette or electronic nicotine product advertisement | 0.854 |
| 7 | X04 PT0021 | Youth has ever used a tobacco product (parent or guardian report) | 0.850 |
| 8 | X04 YC1103 | Ever been curious about smoking a cigarette | 0.850 |
| 9 | X04 YC1104 | Think you will try a cigarette soon | 0.850 |
| 10 | X04 YV1143 | People who are important to you: Their views on using e-cigarettes or other electronic nicotine products | 0.850 |

**Table S1**: Top 10 most important variables from DeepSeek-V3

| **Order** | **Order** | **Variable** | **Variable description** | **Importance Score** |
| --- | --- | --- | --- | --- |
| 1 | X04 YX0681 | X04 YX0681 | How many of your best friends use e-cigarettes | 0.912 |
| 2 | X04 YX0716 | X04 YX0716 | In next 30 days, likely to purchase e-cigarettes or electronic nicotine products | 0.908 |
| 3 | X04 YV1143 | X04 YV1143 | People who are important to you: Their views on using e-cigarettes or other electronic nicotine products | 0.879 |
| 4 | X04 YX0741 02 | X04 YX0741 02 | People who are important to you use: E-cigarettes or other electronic nicotine products | 0.875 |
| 5 | X04 YX0678 | X04 YX0678 | In past 30 days, received a free sample of an e-cigarette or other electronic nicotine product | 0.847 |
| 6 | X04 YV1124 | X04 YV1124 | Likeliness of someone becoming addicted to e-cigarettes or other electronic nicotine products | 0.846 |
| 7 | X04 YV1149 | X04 YV1149 | Harmfulness of nicotine in electronic nicotine products to health | 0.843 |
| 8 | X04 YV1099 | X04 YV1099 | Harmfulness of using e-cigarettes or other electronic nicotine products compared to smoking cigarettes | 0.842 |
| 9 | X04 YV1125 | X04 YV1125 | Thoughts on how much people harm themselves when they use e-cigarettes or other electronic nicotine products | 0.837 |
| 10 | X04 YX0680 | X04 YX0680 | How many of your best friends smoke cigarettes | 0.832 |

**Table S2**: Top 10 most important variables from GPT-4o

| **Order** | **Variable** | **Variable description** | **Importance Score** |
| --- | --- | --- | --- |
| 1 | X04 YX0716 | In next 30 days, likely to purchase e-cigarettes or electronic nicotine products | 0.950 |
| 2 | X04 YV1124 | Likeliness of someone becoming addicted to e-cigarettes or other electronic nicotine products | 0.867 |
| 3 | X04 YX0715 | Level of ease for youth to buy e-cigarettes or other electronic nicotine products | 0.853 |
| 4 | X04 YV1099 | Harmfulness of using e-cigarettes or other electronic nicotine products compared to smoking cigarettes | 0.850 |
| 5 | X04 YV1143 | People who are important to you: Their views on using e-cigarettes or other electronic nicotine products | 0.850 |
| 6 | X04 YX0678 | In past 30 days, received a free sample of an e-cigarette or other electronic nicotine product | 0.850 |
| 7 | X04 YX0681 | How many of your best friends use e-cigarettes | 0.850 |
| 8 | X04 YV1149 | Harmfulness of nicotine in electronic nicotine products to health | 0.843 |
| 9 | X04 YX0741 02 | People who are important to you use: E-cigarettes or other electronic nicotine products | 0.843 |
| 10 | X04 YX0694 | Parents reaction if they found out you used e-cigarettes or other electronic nicotine products | 0.840 |

**Table S3**: Top 10 most important variables from LLaMA 3.1-70B

| **Order** | **Variable** | **Variable description** | **Importance Score** |
| --- | --- | --- | --- |
| 1 | X04 YX0716 | In next 30 days, likely to purchase e-cigarettes or electronic nicotine products | 0.950 |
| 2 | X04 YX0678 | In past 30 days, received a free sample of an e-cigarette or other electronic nicotine product | 0.893 |
| 3 | X04 YX0715 | Level of ease for youth to buy e-cigarettes or other electronic nicotine products | 0.893 |
| 4 | X04 YX0681 | How many of your best friends use e-cigarettes | 0.883 |
| 5 | X04 YX0708 02 | In past 12 months received discounts or coupons: E-cigarettes or other electronic nicotine products | 0.880 |
| 6 | X04 YX0741 02 | People who are important to you use: E-cigarettes or other electronic nicotine products | 0.880 |
| 7 | X04 YV1099 | Harmfulness of using e-cigarettes or other electronic nicotine products compared to smoking cigarettes | 0.853 |
| 8 | X04R Y YX0671 | DERIVED - Recoded anyone who lives with you now uses tobacco (4 levels) | 0.850 |
| 9 | X04 PR1051 | Rules about using e-cigarettes or other electronic nicotine products inside home | 0.850 |
| 10 | X04 PT0021 | Youth has ever used a tobacco product (parent or guardian report) | 0.850 |

**Table S4**: Top 10 most important variables from Qwen 2.5-72B-Instruct

| **Rank** | **Variable** | **Description** |
| --- | --- | --- |
| 1 | X04R Y YX0671 | DERIVED - Recoded anyone who lives with you now uses tobacco (4 levels) |
| 2 | X04 PR1051 | Rules about using e-cigarettes or other electronic nicotine products inside home |
| 3 | X04 PT0029 | Cigarettes or tobacco might be available to youth at parent or guardian’s home |
| 4 | X04 PX0707 | Youth’s close biological relatives have used any tobacco products |
| 5 | X04 YC1104 | Think you will try a cigarette soon |
| 6 | X04 YC1105 | Would smoke a cigarette if one of your best friends offered you one |
| 7 | X04 YC1206 | Think you will smoke a cigarette in the next year |
| 8 | X04 YC9043 | Likely to become addicted to cigarettes |
| 9 | X04 YV1099 | Harmfulness of using e-cigarettes or other electronic nicotine products compared to smoking cigarettes |
| 10 | X04 YV1124 | Likeliness of someone becoming addicted to e-cigarettes or other electronic nicotine products |
| 11 | X04 YV1125 | Thoughts on how much people harm themselves when they use e-cigarettes or other electronic nicotine products |
| 12 | X04 YV1143 | People who are important to you: Their views on using e-cigarettes or other electronic nicotine products |
| 13 | X04 YV1149 | Harmfulness of nicotine in electronic nicotine products to health |
| 14 | X04 YX0012 | General perception: Most people disapprove of e-cigarettes or electronic nicotine products |
| 15 | X04 YX0071 | People who are important to you: Their views on tobacco use in general |
| 16 | X04 YX0200 | Reaction if parent/guardian found you using tobacco |
| 17 | X04 YX0203 02 | In past 30 days, noticed e-cigarettes or other electronic nicotine products being advertised: At gas stations, convenience stores or other retail stores |
| 18 | X04 YX0203 06 | In past 30 days, noticed e-cigarettes or other electronic nicotine products being advertised: On television |
| 19 | X04 YX0203 09 | In past 30 days, noticed e-cigarettes or other electronic nicotine products being advertised: On websites or social media sites |
| 20 | X04 YX0677 09 | In past 30 days, noticed cigarettes or other tobacco products being advertised: On websites or social media sites |
| 21 | X04 YX0678 | In past 30 days, received a free sample of an e-cigarette or other electronic nicotine product |
| 22 | X04 YX0680 | How many of your best friends smoke cigarettes |
| 23 | X04 YX0681 | How many of your best friends use e-cigarettes |
| 24 | X04 YX0694 | Parents reaction if they found out you used e-cigarettes or other electronic nicotine products |
| 25 | X04 YX0708 02 | In past 12 months received discounts or coupons: E-cigarettes or other electronic nicotine products |
| 26 | X04 YX0709 | Has a favorite e-cigarette or electronic nicotine product advertisement |
| 27 | X04 YX0715 | Level of ease for youth to buy e-cigarettes or other electronic nicotine products |
| 28 | X04 YX0716 | In next 30 days, likely to purchase e-cigarettes or electronic nicotine products |
| 29 | X04 YX0741 01 | People who are important to you use: Cigarettes |
| 30 | X04 YX0741 02 | People who are important to you use: E-cigarettes or other electronic nicotine products |
| 31 | X04 YZ1002 | Ever used any other tobacco products |

**Table S5**: Shared top 50 important variables across four distinct LLMs
